# Fear, Stigma and Uncertainty: The short and long-term effects of Ebola on survivors, affected families, and community in Bundibugyo, Western Uganda

**DOI:** 10.1101/2024.11.08.24316967

**Authors:** Shamilah Namusisi, Jacinta Mukulu Waila, Sarah J. Hoffman, Cheryl Roberston, Katey Pelican, Michael Mahero

**Affiliations:** Makerere University, Kampala, Uganda; Heidelberg Institute of Global Health, University of Heidelberg, Germany; School of Nursing, University of Minnesota; Department of Veterinary Population Medicine, University of Minnesota, Saint Paul USA; University of Tennessee College of Veterinary Medicine, ^5^ Department of Veterinary Population Medicine, University of Minnesota, Saint Paul USA

**Author notes:** Corresponding (SN).

**Keywords:** Affected families, Bundibugyo, community, Ebola, ethnography, survivors

## Abstract

In 2014, Uganda was identified as a high-risk country for Ebola Virus Disease (EVD), with a series of outbreaks recorded since 2000. In 2007, the second outbreak in Bundibugyo district resulted in 149 reported cases and 37 confirmed Ebola deaths. Through the outbreak response, a new strain of the Ebola virus (*Bundibugyo ebolavirus)* was discovered. Although much is known about the nature of Ebola, including disease signs and symptoms, transmission and management, there is limited understanding of the short and long-term sociocultural impacts of the disease in communities. The study team conducted a focused ethnography in Bundibugyo District 10 years after the 2007 outbreak. Data collection included a review of archival data, participant observation, field notes and 19 in-depth interviews with survivors and affected families. Results underscored time-limited social, cultural and economic disruptions caused by the outbreak. We interpreted findings through an Eco-health framework with an emphasis on ways that underlying stigma accentuated detrimental long-term effects of the outbreak. Affected women, particularly widows, experienced social exclusion, and economic strain, and acknowledged loss of opportunity for a better life for their orphaned children. Deepening fear of the possible recurrence of Ebola resulted in ethnic tension driven by speculations on the 2007 outbreak source. Survivors reported varying persistent health effects including impaired vision and general body weakness. Community members reported positive changes in health seeking behaviors. Health care workers described high levels of alert for early clinical signs of Ebola, a critical factor for early outbreak detection at the community level. Our findings can inform future Ebola response and recovery interventions, particularly those targeting community re-integration and a mitigation of the fear and stigma associated with survivorship.

## Introduction

Since the identification of the Ebola virus in 1976, Africa has suffered the highest number of outbreaks globally [1]. It is generally estimated that the number of outbreaks are going to increase across Sub-Saharan Africa as habitats for Ebola aggregate [2]. Over the last two decades, two major outbreaks have been recorded, in West Africa (2013-2016) the largest and deadliest, and in Eastern part of Democratic Repulic of Congo (DRC), causing 11.310 cases and 2,287 deaths [3,4] respectively. The outbreak in DRC is believed to be the second largest and most complicated to date due to conflict, political instability, poor infrastructure and socio-economic weaknesses [3,5,6]. Ebola disproportionately impacts people living in rural communities in low income countries causing disruption of various economic activities [7,8]. The origin of Ebola outbreaks in Africa has tended to be rural and among impoverished communities, including the West Africa outbreak that started in rural Guinea and expanded to the level of an epidemic [9–11]. Ebola places a burden on communities and nations through loss of money and economic activities and capital, an increase in unemployment and brain drain, slowdown or closure of health facilities, disruption of internal and external trade, decrease in tourism and travel activity, and an increase in social and cultural barriers [3,11].

EVD outbreaks affect many aspects of life at individual, household and community levels [12]. The outbreak in West Africa further highlights the outbreak’s impact on the population, health care system and economy of affected countries [13]. Previous studies primarily focused on the disease’s spread, clinical signs, diagnostics, prevention, and more recently, vaccine and drug development [14]. However, this alone cannot comprehensively explain the devastating consequences the disease has had in the lives of those diagnosed with EVD and their families [15,16]. Studies of mental health effects in West Africa have found high prevalence of symptoms related to depression, anxiety and psychological distress due to stigma [12,17].

The stigmatization of survivors, affects the ability to establish effective response strategies following an outbreak [18]. Disease-related stigma and its sequalae have been documented previously in the context of EVD. The 2000-2001 outbreak in the Gulu, Masindi, and Mbarara districts of Uganda resulted in social rejection, harassment, and discrimination of Ebola survivors [19]. During the 1995 EVD outbreak in Kikwit, Zaire (renamed Democratic Republic of the Congo), EVD survivors returned to their communities’ post-recovery to find their homes and belongings had been burned, and many times spouses had fled [20]. Community members refused to buy market goods from EVD survivors post-return, influencing economic status and position in the societal hierarchy [21]. The sociocultural effects of disease-related stigma can have dramatic effects on housing, criminal activity, meaningful relationships, and individual wellbeing [3,22]. These effects also drive larger, collective factors such as food insecurity, displacement and environmental degradation, which are directly relevant to human interaction with the environment [22].

Uganda has experienced several outbreaks of Ebola virus disease (EVD) and the most recent outbreak in Uganda started in September 2022 in Mubende district, spreading to other neighboring districts [23,24]. The country’s first and largest outbreak occurred in the Northern district of Gulu in 2000 [25,26]. This was followed by several other outbreaks: 2007 in Bundibugyo district, 2011 in Luwero (single case, one death) and the 2012 outbreak in Kibale [27]. On 29 November 2007, the Uganda Ministry of Health (MoH) and the World Health Organization (WHO) confirmed an outbreak of EHF in Bundibugyo District, western Uganda and On 20 February 2008, the outbreak concluded with 93 putative and 56 laboratory-confirmed cases[28]. There were 37 case fatalities in an outbreak that saw the discovery of a new strain of Ebola (*Bundibugyo ebolavirus)* (CDC, 2007). Notably, the 2012 outbreak in Isiro, Democratic Republic of Congo was retrospectively attributed to the same strain *Bundibugyo ebolavirus* [28,29]. The majority of cases (36%) in the Bundibugyo outbreak were subsistence farmers and 12% were health workers [30]).

A characterization of short and long term impacts of Ebola outbreaks is a more recent area of focus in research [13,17,31,32]. Apart from the immediate health effects, the disease also instigates a breakdown in social and cultural values or norms that have long been known as major survival strategies among resource-limited communities such as those in Bundibugyo [3,26,29] . Survivors and affected communities experience short and long-term consequences of Ebola [12,32]. The short-term psychological effects among survivors include the traumatic disease course and the physiological trauma associated with fear of death and proximity to others dying [27,30]. Survivors report feelings of shame or guilt and stigma or blame from others [29,33].These are compounded by a change in cultural practices surrounding many of the traditions that long held communities together [16,29]. At the community level, there is an immediate sense of grief due to loss of community members and shifting family structures due to loss of parents, primary income earners, health workers, community leaders and other social services provision professionals[34]. The United Nations Development Program (2014) predicted an orphan crisis and disruption of longstanding traditional community support in West Africa caused by the Ebola outbreak [8]. However, no parallel studies describe the Ugandan context.

## Purpose

The aim of this study was to understand socio-economic and cultural effects of Ebola among survivors, affected families and communities post-outbreak in Bundibugyo district, Uganda. The findings are important in the design of recovery programs targeting Ebola affected individuals and communities. Moreover, our findings add to the emerging body of literature describing the short and long-term impacts of epidemic/pandemic courses in affected communities.

## Conceptual Framework

We used an Eco-health approach integrated with perspectives on disease-related stigma to design the study and interpret reported individual, familial, and community experiences on EVD survivorship in the ten years since the Bundibugyo outbreak. In studies involving zoonotic diseases, where there are fundamental connections between human, health, and environmental systems, examining factors that potentiate vulnerable points in these relationships offers a greater understanding of the phenomenon as well as potential mechanisms for intervention (Ecohealth). Based on our work and the work of others in the social, cultural, and economic intersections of zoonoses, we framed our analysis through the perspective of Ecohealth and stigma. Stigma is a collective, negative social reaction to an individual attribute that is specific to time and place (Miles, 1981). Depending on contextual factors, enduring disease-related stigma is associated with social rejection, violence, long term inequities in the distribution of income, housing, and health, and significant, negative psychosocial effects of survivorship [32,34,35]. In the context of an outbreak, stigma and resulting discrimination are potentially more damaging than the pathology of the disease itself [34].

## METHODS

### Study area

This study was conducted in Bundibugyo district and data were collected over a period of three months from October to December 2015. The district lies in western Uganda bordering the DRC and is the only district west of the Rwenzori Mountains. It is bordered by Ntoroko District to the northeast, Kibaale District to the east, Kabarole District to the south, and the DRC to the west and north. The district headquarters at Bundibugyo town council are located approximately 32 kilometers (20 miles), by road, west of Fort Portal. Though part of the Nile basin, Bundibugyo district is ecologically and culturally part of Central Africa with similar customs to those of eastern DRC. The study took place in Bundibugyo town council and Kikyo, about 25 kilometers apart. Kikyo, the epicenter of the outbreak, is characterized geographically by low lying and mountainous areas rising to about 6200-8900ft above sea level. Its rainfall distribution enables agriculture to take place throughout the year. It is also surrounded by swamps, rivers like Humya, Kirumya forest (Semuliki) that harbors wild species like monkeys, chimpanzees, buffalos among others. The communities adjacent to the forest practice subsistence agriculture and use the forests to supplement their livelihoods. Some of the forest products include bush meat, herbal medicines, fruits, vegetables, fuel in form of firewood and charcoal, and construction materials such as timber and vines for making ropes. Therefore, the forest is of great social-economic importance to the local communities. Land for agricultural production is steadily shrinking due to an increase in population and the fact that this area is sandwiched between the Semuliki national park and North Rwenzori Forest Reserve.

## Map showing Ebola Outbreak Areas in Uganda

### Study design

We conducted a focused ethnographic inquiry collecting data through: in-depth interviews with survivors, affected families and community members, participant observation, extensive field note documentation and archival data review. The study team consisted of five scholars with veterinary and nursing backgrounds led by an expert in ethnographic research methods.

### Participant sampling

Initially, purposive sampling and subsequently snowball sampling techniques were used to identify interview participants. All study participants were 18 years and older and included survivors and affected families and community members that were residents in the area at the time of the outbreak. Survivors were adults who self-reported laboratory confirmed EVD infection, and who were hospitalized and discharged after laboratory confirmed negative status. Affected families were those who had a member suffer from EVD regardless of whether they survived or succumbed to the disease. The community refers to residents of the study area who were present at the time of the outbreak.

## Data collection procedures

### In-depth interviews

Interviews were conducted either in English or local languages with the aid of an interpreter who was a trained qualitative research assistant and a native of the area, fluent in Rubwisi and Rukonzo (the two major local languages in Bundibugyo) and highly proficient in English. An in-depth interview guide provided the interview framework. All interviews were audio recorded with participant consent and written notes were taken during the interviews. A total of 19 interviews were conducted.

### Archival data

The team reviewed medical records of patients admitted with Ebola at Bundibugyo hospital. These records showed who was affected, the area of residence, ethnicity, religious affiliation, sex, age, dates of admission. Treatment received and whether recovered or died

### Participant observation

Data collectors recorded community observation, including activities being carried out, the general home environment, how people interacted with each other, and key issues emerging out of the conversations besides the interviews. All data was collected between November 1 and December 20, 2015.

### Data management

Verbatim transcription of the audio files was performed and the interviews that were conducted in local languages translated into English. Investigators reviewed the transcripts with the study interpreter after the translation was complete. Independent verification of all translated transcripts was done by a second study team member who was not involved in the original translation and fluent in one of the local languages and English. Finally, all transcripts were subjected to quality checks against the audio recordings and compared with the hand written notes by the translator and the investigators.

### Data Analysis

Thematic qualitative analysis was performed in close collaboration with the research assistant who did interpretation during data collection. Two researchers with expertise in qualitative methods independently read at least six transcripts and developed a list of codes through open descriptive coding. The codebooks were then merged into a single codebook through review, discussion, and consensus. The codebook was then applied to manually analyze the data using Microsoft Word. Analysts constructed a table that included all codes where excerpts of texts from the transcripts that corresponded to the codes were copied and pasted into the table. After coding seven transcripts together revising the codebook iteratively, the two analysts then divided the 12 remaining transcripts equally to complete coding, meeting after every three transcripts to review and integrate new codes into the coding frame and back code when appropriate. Once coding was complete, the codes were then grouped together into categories guided by similarity in content after which the categories were further collapsed into themes. The field notes and archival data were used during data analysis to give context to participants’ narrations, aiding with interpretation.

Trustworthiness was established through analytic procedures, that included maintenance of an audit trail of audio files, close collaboration with a consistent study interpreter, memoing, and other methodological procedures like field note review, rich description of participant observation, and the cross-referencing of findings across the categories of respondents

### Funding information

The study was conducted with funding support from University of Minnesota as a capacity building grant for the then One Health residents under the USAID first funded program of Emerging Pandemic Threats (EPT1).

### Ethical considerations

We received ethical approval to conduct this study from the Joint Clinical Research Center in Uganda and the University of Minnesota Institutional Review Board (IRB Code Number: 1502S62201). Written informed consent was obtained from all study participants. Additionally, Bundibugyo local government, through the office of Chief administrative officer granted the study team permission for community entry. The medical superintendent approved access to medical records and this facilitated the recruitment of study participants.

## Findings

### Characteristics of study participants

Table 1 describes the characteristics of the key informants. The age of participants ranged between 32 and 52 years with the mean and median being 43 years. Majority of the participants were male (74%) while females were about a quarter (26%). All participants reported attaining some level of education ranging from primary school to graduate levels. Whereas men engaged in various income generating activities, all women were engaged with unpaid work in the home. Among male participants, paid employment varied by education level whereby college graduates worked as district technical leaders (16%), health workers (16%) and social workers (11%). Most of those educated below post-secondary level worked as farmers (32%). All participants reported being affiliated to various religious groups with the majority (90%) identifying as Seventh Day Adventists (SDA).

**Table 1.**
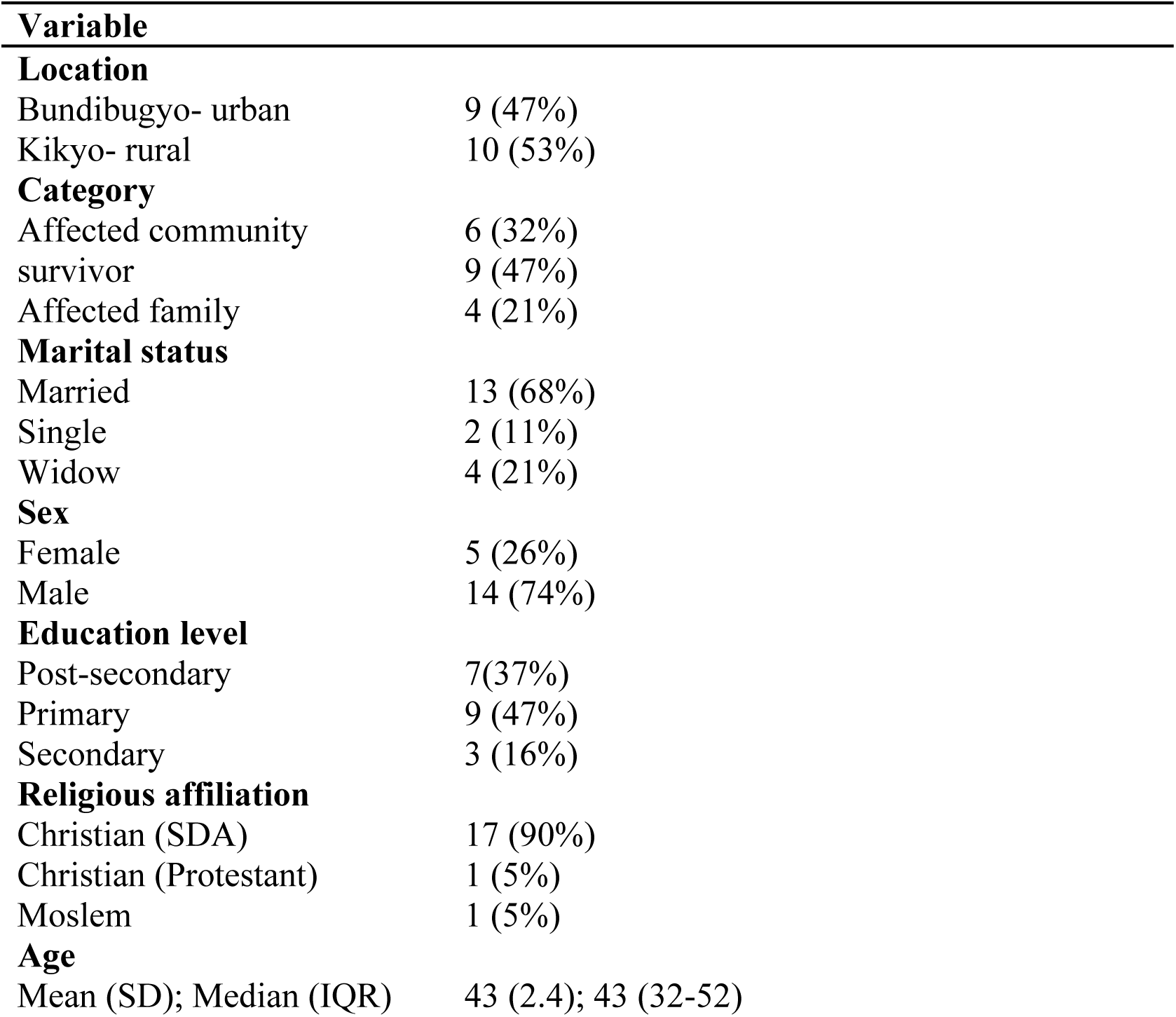
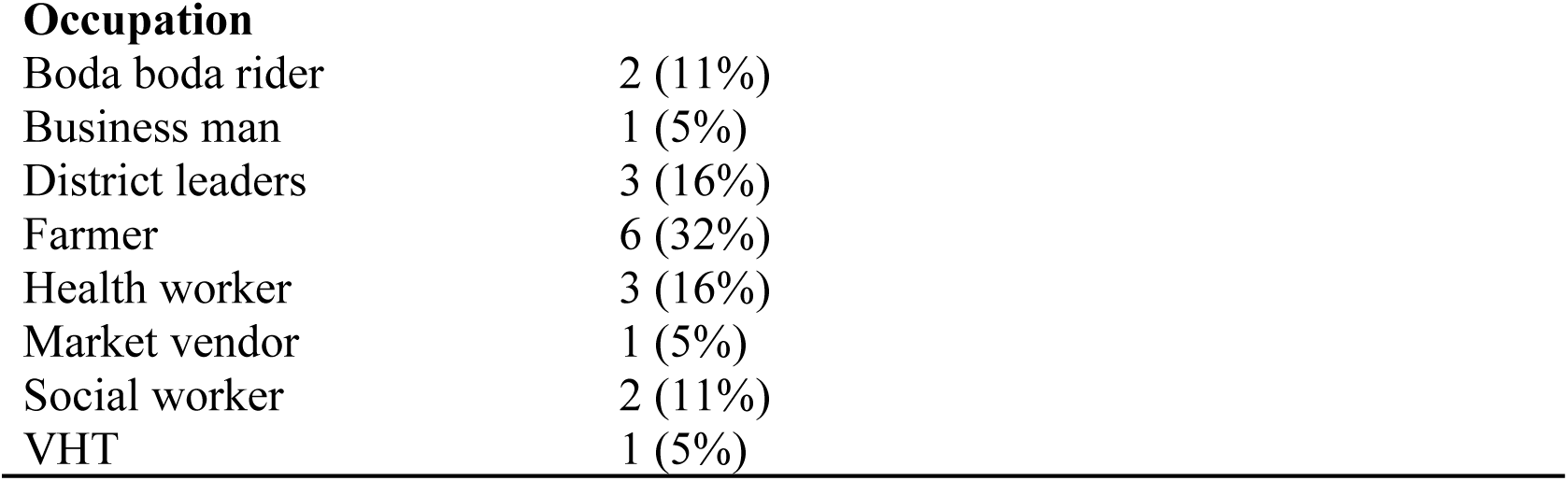
Summary of respondents Descriptive statistics (n=19)

### Description of general life before the Ebola outbreak

Participants described their lives before the outbreak as being “normal”, men and women went about their daily routine activities of providing for the families and caring for the children respectively. Community gatherings went on without any restrictions (freedom of association), children went to school and non-governmental organizations were present to offer humanitarian aid to address various challenges in the community. Cocoa farming, the main cash crop in the area, was the major economic activity undertaken by most residents. The area faced the challenge of poor road infrastructure and the main road to Bundibugyo from the neighboring districts was as poor as the road network within the district. Bundibugyo district hospital was the main health facility serving the population supported by other health facilities of lower levels, including Kikyo health center III, where the first Ebola cases were reported.

### Emerging themes

Three major themes emerged during data analysis; i) The Ebola outbreak, ii) The effects of the outbreak, and iii) Life after the outbreak, as described below. Quotes are identified using the interview number and the category of the participant (survivor, affected family or community member). The acronym ‘CHW’ is used for Community Health Workers, ‘HSP’ is used for health service providers such as nurses, clinical officers, and medical doctors.

### Theme 1: The outbreak

This theme presents the community experiences during the Ebola outbreak from the detection of the viral disease to its containment. It has two main subthemes:

**Subtheme 1a: The health care system response** including the challenges with diagnosis and management of an unusual health situation are presented under this subtheme.

**The Strange Disease:** Around August 2007, participants reported that community members began falling ill from what was described as a strange disease. To the health workers and the community members alike, this strange disease has signs and symptoms like those of malaria and was managed as such.

> *“The clinical officer who was very instrumental in diagnosing…had some disease that he called strange of course…A fever, diarrhea and vomiting so anyone would have thought that this was possibly what? Malaria. So he started I think managing, managing, managing but at the end people were not improving.” - IDI002-HSP “Ebola started with our dad and it was the first family to get affected. We first heard of it in a place called Kikyo. That malaria has come to Kikyo*.”-IDI004-Survivor&HSP

To bring the situation under control, health workers exchanged information among themselves, although EVD suspicion was very low partly because none of the health workers had prior clinical experience with such a disease.

> *“When the disease occurred in Kikyo for 2 months…a nurse in Kikyo called me and explained to me what was happening and I felt that it could be a Hemorrhagic Fever (HF).*” – IDI001-HSP

The characteristic symptom of bleeding from body openings was only observed in a few of the cases, further eluding the suspicion of hemorrhagic fever.

> *“People had no knowledge about Ebola. Of course, people had heard…during the Gulu outbreak but had never had knowledge to know how it spreads, how it looks, of course we used to hear may be on news on how it spreads but being in Gulu which is far away from Bundibugyo*.” **–** IDI003-NGO worker
>
> “………the way it came was not really not typical…people will start thinking of Ebola when you have bleeding from anywhere…the kind of presentation that was here that would possibly be the latest sign.” - IDI002-HSP

Health care workers at Kikyo health center, the epicenter, reported cases to the district and the Ministry of Health in Kampala, appealing for support to diagnose the strange disease.

> *“…People are not improving…at a certain point, the district reported, even officials from the ministry came, they went to Kikyo down at the ground and tried to investigate, took samples and things of that kind*.” – IDI002-HSP

### Complicating factors for the outbreak

Some participants expressed concerns that the imminent hosting of the Commonwealth Heads of Government meeting of November, 2007 by Uganda somehow delayed the detection and declaration of the outbreak. Similarly, during that same time, the district had an ongoing typhoid fever outbreak which masked the fact that something new was coming up.

> *“So, within that period of typhoid fever also Ebola came in so the whole thing was just happening …. I think at one time some of those patients ended in theater thinking that maybe they have gotten perforated gut and they were opened up maybe to find inside there is nothing happening.”* **–** IDI002-HSP

### Ebola confirmation, fear and panic during response

Amidst the confusion of a strange disease with associated death, the Ministry of Health sent a technical team to assess the situation and samples were taken which finally confirmed it was Ebola.

> *”It began in August in Kikyo and started getting cases in the last week of September and managed cases in the hospital until 30th October when it was announced as Ebola*.” IDI001-HSP

Having heard about the devastating nature of Ebola from the Gulu outbreak, the declaration that the strange disease was Ebola sent waves of panic across the entire community. Health workers experienced as much fear as lay community members and this fear led to disruption of healthcare service delivery.

> *“Most of the staff in the ward and even patients fled due to fear…… That experience was really terrible for me and my colleague. Even my family members often pestered me to leave unit/hospital but I told them no. What I know is that God kept me for a purpose and He knows it. The entire time I did not step home until Bundibugyo was declared Ebola-free*.” – IDI001-HSP
>
> “*The professional health workers had fled the hospital but for us as volunteers we would not leave our community. – IDI018-CHW, boda boda rider & burial team member*

Similarly, other social and economic activities were also affected:

> *“The bank was the only way to get…support in terms of funds to run these activities. But you are going to the bank, the bank manager, everybody in the bank they are scared*.” – IDI003-NGO worker
>
> *“So indeed, there was stampede so to say…every person packed and left the district including even government workers those who were not from here. They packed and left.”* – IDI014-Government official

### Subtheme 1b: The affected communities’ response

This subtheme expounds on how local community members reacted to the emergence of a strange disease which the healthcare system was unable to diagnose.

### Suspected witchcraft

With no clarity on what was happening, community members speculated to explain the unusual occurrences. For some families, it was an act of witchcraft. The concern was, why people from the same family experiencing the same signs? If the problem was medical, why were medical workers failing to diagnose and or treat the disease?

> *“Partly the witchcraft issue came before the declaration was made, people were not aware of what was killing*.” – IDI003-NGO worker
>
> “*People were thinking that maybe there are some charms and there was a story somewhere that someone there is a witch doctor so people were thinking that maybe because he was getting some people from Congo coming for those herbal medicines*.”-IDI008-Survivor &HSP

### Act of sacrifice

In one of the villages where six people died in a family, community members thought that it was an act of sacrifice by one young man, a member of the same village, to get richer. At one of the burials, he was attacked and almost beaten to death causing him and his family to migrate to Kasese district.

> *“One home in Butholya six people died at once. There was a young rich man in that village, they thought he is the one sacrificing them, even on the burial people had to beat him. It was after the death of the 5th person and people thought that maybe he is getting……he wanted to get money using “mayembe” (local charms), up to now that man shifted to Kasese and said he will never come back, he hates the whole clan and the family.”-* IDI018-HSP

### Suspected poisoning and tribal conflict

Though tribal tensions between the Bakonzo and Bamba ethnic groups in this area existed long before the Ebola outbreak, the confusion surrounding ‘the strange disease’ seemed to have rekindled the rift between the two tribes. Most of the initial cases of ‘the strange disease’ were reported in mountainous areas among the Bakonzo. There emerged a confusing narrative that the Bamba who live at the foot of the mountain had poisoned the water source causing death of the Bakonzo. This claim resolved after ‘the strange disease’ was confirmed to be Ebola.

> *“I remember at first there was general complaint that there is a strange disease and it is killing people. You know here we having majorly like two or three tribes; the Bakonzo, the Bamba, the Babwisi and this strange disease was in the area dominated by the Bakonzo. So, I remember they came to our office, the CAO’s office complaining that, the Bakonzo have been poisoned, you know*.” – IDI014-Government official

### Acceptance and compliance

Following the official declaration of an Ebola outbreak, community attention shifted to a positive response to an outbreak situation, highly influenced by what they had heard about previous Ebola outbreaks. The immediate reaction was that of acceptance and compliance to the situation and fear for one’s self.

> *“We’ve heard of Ebola in Sudan; we had heard of Ebola in Congo so the moment people heard this is Ebola there was no one like to contest it. People knew this was Ebola the only problem now was everyone was starting to fear for him/herself”* **–** IDI002-HSP

Community members exhibited heightened levels of suspicion which motivated cooperation with the outbreak response teams evidenced by reporting suspected cases to health workers to facilitate prompt isolation and treatment.

> *“……he used to work in the taxi park, he vomited in the park and since vomiting was one of the major signs of Ebola and everyone knew it, they ran away. Now that there was this surveillance and mobile team with ambulances ever ready, they came for him and he was running because he knew that he had no Ebola, he had just taken a lot of waragi(local alcohol drink) that’s why he vomited*.” – IDI019-Affected family

### Use of dangerous remedies

Communities adopted various strategies to ensure their safety including excessive use of chemicals like sodium hypochlorite (JIK) causing hand burns to some and abdominal pains among those who attempted to drink it.

> *“People were even starting to take JIK thinking it is something that can, you know people were saying, now if you take JIK you… so people were even washing hands with JIK and people got some burns.”* **–** IDI003-NGO worker

Additionally, people resorted to high consumption of *waragi*, a local alcoholic drink. The assumption was that the alcohol would cleanse the system off the “Ebola germs”. High consumption of waragi was also supported by the community observation that some of the people who were recovering from the disease, were those that practiced daily consumption of alcohol.

> *“There were three beliefs (about the relationship between waragi and Ebola); if someone was a drunkard, he could not die of Ebola, because it’s only those who had never tasted alcohol that died and only those who were known drunkards survived. Waragi being a spirit helps to clear the stomach and Ebola was associated with taking dirty things. and lastly that drinking makes people not to move, so you stay in the same place reducing the risk of getting outside.”* **–** IDI019-Affected family

### Theme 2: The effects of the outbreak

The outbreak disrupted normal activities, amplifying fear, and stigmatizing affected individuals and their families. Media messages, the exodus of non-locals, and the shunning of people from Bundibugyo wherever they went heightened this stigma. In efforts to control the outbreak, various regulations were implemented, impacting life in multiple spheres and to varying degrees. This theme explores the short-term effects of the Ebola outbreak on the community at different levels

### Subtheme 2a: At the social level

Various containment measures that were put in place affected the social lives of the community members.

**Social gatherings’ restriction** meant ceremonies like weddings and prayer gatherings were completely put off:

> *“Completely no, you couldn’t do a wedding. Those are the things you could not talk about. What was here was basically fighting the disease and looking for what to eat. These other social issues, we would not talk about them*”– IDI014-Government official
>
> “*Well, I should say of course when something like this was declared, then the local government, the central government, the political wing of course they put lots of regulations like they already started restricting gatherings. I can say people started shying going to church because when in church you are going to sit together, shake hands and so some people started shunning some of these functions*” – IDI002-HSP

### Rejection by community members

Those who came into contact with or were associated with Ebola patients faced rejection from the rest of the community. Transport operators, especially *boda boda* riders who transported Ebola patients and the bodies of victims, bore the brunt of this rejection

> *“We used to carry patients, but we didn’t know which disease they were suffering from. Then people started dying, only to hear that its Ebola. One day one of the patients died and everyone was fearing to carry the body. For me I decided I would do it after all the body was already in the coffin. My other friend also did the same on a different day and then people started fearing us that those ones carry Ebola, saying that’s the one who carried Ebola he is going to affect us, they would run away whenever they would see us at a distance. We used to go to restaurants and they would refuse to give us food because we carried bola patients and dead bodies*.” **–** IDI015-Boda boda rider

**Stigma and trauma** suffered by Ebola survivors and their families driven by fear among community members that survivors were still infectious.

> *“We came back to our home in town…It took long time for people to get used to, before they could come near us to even touch us. You know people before they trust that you don’t have the disease, it takes a person time about a month or two. We spent about two months in the community to fit in the community*.” – IDI004-Survivor
>
> *“There was a day I came to town to cut my hair and when the people scampered away saying I am an Ebola patient. People used to run away from us*.” – IDI006-Survivor
>
> *“But people feared us, whenever they could see us at a distance, they would run away [laughing]….it was a bad period for me*.” – IDI010-Survivor

A breastfeeding mother described what she and her baby went through when she returned from hospital after surviving Ebola

> *“It was the father who used to take care of the baby. I did not have anyone else to take care of me. He used to carry the baby while putting on gloves. He would take the baby and bring him at a time of breast feeding. My husband had to take that risk as a parent; he couldn’t throw me away* **–** IDI017-Survivor

### Subtheme 2b: Cultural changes

Burial ceremonies and funeral rites suffered major disruptions.

Death and grief were experienced by the entire community but the effects were more at the family level with some losing several of their members to Ebola.

> *“Very many people died. You know where there is death, it is a disaster…some families where both a father and a mother died, some families a father died, some families a mother died, so as you know where there is death there automatically must be some challenges which are negative to the lives and of course being that the end of it all who are the victims of such a situation? There are the children. Most children were left orphan and now they had no people to look after them.”* **–** IDI003-NGO worker

There were no children reported to have caught or died from the disease even though one woman continued to breastfeed when she was suffering from Ebola and after recovery.

> *“They used to bring the baby in the evening, and we sleep together, and they come for him very early in the morning and baby didn’t get the disease. I would breast feed him very early in the morning then they take him the whole day. They would give him porridge and milk during daytime. He was like 3 months old that’s why I had to breast feed him*.” – IDI017-Survivor

### Burial teams and sites

Most families were given the opportunity to bury their dead in their homes under the supervision of burial teams except for a few categories of people like health workers and the religious leaders who were buried at the hospital and churches respectively. However, since the usual burial ritual were not conducted, their loved ones were left devastated.

> *“He (my husband) was not buried in the normal way, they did not wash him, they just brought him from the hospital after they had wrapped and put him in the coffin direct into the grave, they did not perform any rituals. Too bad but there was nothing to do, it was touching, because I could not burry him in the normal way.” **-** IDI005-Survivor*
>
> *“You know when someone dies here there are so many practices, you wash the body, you want to bury your person, you want to mourn him very grievously but all those things because laws were in place, they changed ideally because like in this case no family was allowed to bury its own person. This is against culture but I think people allowed, people accepted because at the end people heeded to what the government had said and started allowing their people to be buried by the authorities so ideally so many things changed. You cannot just expect it to be like that that your person dies and you just give the corpse to other people to go and bury for you that’s not acceptable in cultures but they accepted and so they allowed the burial teams to move around looking at who has died, they collect, either if you didn’t they of course meant decisions.”* **-** IDI002-HSP

For those who came from the hills, communities in the lowlands offered land for burial to avoid carrying dead bodies for long distances without motorized transport which would increase the chances of disease transmission.

> *“People were scared. Others would be buried on the way if a vehicle can’t reach your home. They would request any relative who has land where a vehicle can reach to bury there. People would feel bad but there was nothing to do, because we feared the disease, no one could be blamed*.” – IDI016-Survivor

### Hunting practices

Another major cultural shift was suspension of the hunting because of the widely disseminated message that EVD was associated with hunting of wild animals like monkeys and gorillas. Overall, these cultural shifts were temporary, with people quick to reverting to their usual practices once the outbreak was declared over.

> “*People forget easily, our communities forget easily, because if somebody must have eaten a monkey, I should say at this time you cannot stop him if he is still living, if he is a hunter, he is still hunting…Even the animals in the bush were like, why are we not being hunted this time because there was negativity because of the constant information people had to change their way of living.”* – IDI003-NGO worker

### Subtheme 2c: At family level

Interactions among family members were restricted more especially among Ebola survivors and persons who were high risk such as healthcare providers.

### Being cut off from families

This especially affected healthcare workers in the frontline of case management. For some people, they also lost friends and significant others due to the outbreak.

> *“I was cut off from the family because now as I recovered the third week I joined the isolation. I couldn’t even go home I was just at the isolation site. Then after that aha 21 days they monitored me at least that one I went home*” **–** IDI008-Survivor&HSP

Men in polygamous family set ups also stuck to one wife as a means of protecting their loved ones.

> *“I told you that I have two wives. At the time of Ebola I stayed with one woman, I didn’t go to the other wife’s home. I first stayed with one wife & the children were young. I stayed with the wife with the young children. I stayed there until the end of three months that when I went to the other home.”* **–** IDI006-Survivor
>
> *“No. strictly I had stopped sleeping at my homes. I think it took around 4 to 5 months. I could not. But I was with one of my wives here and my children.”* – IDI014-Government official

Sexual restrictions among couples and in cases where one of the partners was a survivor, was another effect that families had to deal with.

> *“There was a little bit challenge came on the issue sex part where of course we would encourage partners to take a little bit long for 2 months, not having sex, Then, they would say, oh! If now you say we should take this long without having sex that means that the person is still dangerous, so that was also again a little bit challenging*” – IDI003-NGO worker
>
> *They said that I had to spend three months without sleeping with my wife. I was put in a separate room. So from there I was give some condoms so that if I feel that the situation was unbearable, I should use a condom*. **–** IDI006-Survivor

### Family breakups/abandonment

In some cases, people separated from their spouses. One *boda boda* rider explained that one of his friends died of Ebola at Bundibugyo district hospital, and because the body was already in the coffin, he decided to carry the body home, something that his wife did not take lightly.

> *“My own wife run away. I decided to leave her because you go when am not sick, what will happen if I became sick? It means you can’t take care of me, we had to separate. I have never even got another one I decided to leave women, that woman made me leave women.”***–** IDI015-Boda boda rider

### Subtheme 2d: Economic disruptions

The economic disruptions affected production, sale and consumption of goods and services.

### Cocoa cash crop

Generally, Bundibugyo is a cocoa growing district and many households depend on the crop for income. Harvesting of the crop usually takes place around October / November. This was the time the outbreak took place, disrupting business activities. The district was cut off even from the neighboring one, where not only were Bundibugyo residents shunned or denied entry, but trade in produce and general merchandize was severely restricted, despite the need for resources to control the outbreak

> *“The economy… there was no business, everywhere business came to a standstill of course who would buy? People were just at home, everybody was scared. You know people would even think that by merely moving, one would probably contract Ebola, but also once you move a head of household would tell you, whoever moves should not come back here, just stay there, so there was no movement, no business.”* – IDI003-NGO worker
>
> *It came at a time when you know this place has a major cash crop called cocoa and there is a season actually that is when it begins from around September, October, November, December, January that’s when the season is peaking.* – IDI002-HSP

### Local businesses/Retailers

At the family level, economic effects varied depending on gender, occupation, role in the community, family size, and infection status. Men, who traditionally are the bread winners, who worked in the business and transport were most affected. One businessman who survived Ebola narrated how difficult it was for him to look after his family during and after the outbreak.

> *“My customers first feared me. It took me about six months and my business was really slow. People used to say they fear me, I still have Ebola….It (business) was closed because if my wife had opened, they would not have given her money, she was also feared & all my people at that time. They feared my family as well…Almost if we could add the months that I spent in the hospital without working, plus the time I spent home, it’s almost three years. The three years were difficult.”* **–** IDI006-Survivor

### Theme 3: Life after the outbreak

This theme presents how the affected community has readjusted to life after the disruptions associated with the outbreak including long-term effects of the outbreak.

### Subtheme 3a: Deepening tribal conflict

The two main communities, the Bakonzo and Bamba, still traded blame long after the outbreak.

> *“Okay, as I talk even now there still those clashes on small scale between the Bakonzo and the Bamba. Other people claim the district is theirs, others claim to be from Kasese so there is no real rich understanding of the point”*.-IDI008-Survivor&HSP

### Insults linked to bush meat consumption

The Bamba ethnic group still believed that the rival Bakonzo tribe who live in the mountains were responsible for the Ebola outbreak and its associated devastation.

> *“The ethnic group which was more affected were Bakonzo…After the ministry of health declared some causes of Ebola disease that it was caused by eating some monkeys and …because of neighboring the Rwenzori mountains where the monkeys stay that it was due to that, they are hunters that’s why they were affected, such a kind of statements are painful*– IDI012-CHW

### Subtheme 3b: The road to recovery

The recovery process varied from one individual to another and from one family to another and was determined by whether the person was a survivor, affected family or community.

> *“The widows still manifest some kind of sadness. Especially one widow has never come back to the normal life. When you look at her she still has some sadness. She is somebody who was depending on her husband and she explains that she has lost that relationship and her way of life has not come back to normal.”*. – IDI001-HSP

### Psychosocial support

To support recovery, immediate psychosocial and material support was provided to the survivors and affected families. Efforts were made, through open discussions, to facilitate understanding, the value of supporting survivors, and that survivors did not possess any risk of spreading the virus. Once the communities understood this, they embraced survivors, a factor that contributed to the reintegration and recovery.

> *“People started knowing that although it is a deadly disease, we can manage. Each one had different feelings. Especially those who lost relatives were grieved for a long time and wondered if they would be compensated. The hardest thing was the psychological trauma… they were very sad and lived with no hope for a long time.”* – IDI001-HSP
>
> *“In the first days and weeks, some of the families feared their fellows after discharge and it took time for them to accept or be in contact with them. After some time, they realized that these people were not contagious. In fact, if you go to the affected homes, you wouldn’t know them not unless they tell you.”* – IDI001-HSP

However, despite the support, some families felt they had not been compensated while some survivors suffered prejudice:

> *“I have been getting some complaints from affected families especially survivors saying that they have not been compensated. The government compensated the health workers only. Their properties were burnt after discharge and communities complain that they were not compensated.”* – IDI001-HSP
>
> *“We lost him (husband) … we are suffering, I am suffering, I am taking care of myself and the kids and I don’t have any support.”* **–** IDI009-Affected family
>
> *“My daughter left me with her two school-going children and we did not get any assistance. I am weak”.* **–** IDI005 -Survivor

### Chronic health conditions

For the survivors, the recovery process was complicated by the trauma they faced while in the isolation ward.

> *“I just knew that I am already dead. You can imagine another patient dying when you are just seeing and you are suffering from the same disease! We were not allowed to get out of the ward, they would keep it closed all the time. I couldn’t think that I can survive*.”IDI016-Survivor
>
> *“Very many people died…you are seeing and another, then another…. the whites would come and wrap the body with buvera (polythene bags), spray and they take the body out.”*– IDI004-Survivor

Some of the other effects described by survivors include general body weakness, headache, hypertension, pneumonia, diabetes, and impaired vision.

> *“The way I see since I got hypertension & diabetes because of too much thinking…I used to think that am dead, am no longer among the live ones. That’s how I think Ebola changed my life. Otherwise, am okay.”* – IDI006-Survivor
>
> *“I have some eye problems, in the morning hours I don’t read very well, that was a after the disease, so I use some glasses to read properly. Then periodically I normally get, we call it lobe pneumonia, its normally one side so that one comes at least every month at the beginning of the month I experience that thing.”* – IDI008-Survivor&HSP

**Improved emergency preparedness and health-seeking behavior** whereby healthcare workers adhere to infection control measures and community members are keen to seek medical attention for unknown or suspicious health conditions.

> *“Before Ebola, some health workers took the use of gloves for granted but now they know the importance of hand washing and use of protective equipment. We have some protective gadgets in the store awaiting any events. Before Ebola we had traditional healers, even now they are there. Some of the patients were rushed to the traditional healers. Whereas they are there, when the community suspects that the disease is unknown, they bring the patient to the hospital.”*-IDI001-HSP
>
> *“So on the basis of this Ebola outbreak, there are so many people who went to study and they are going to come back. I know even of recent there is someone who we just connected to go and study medicine and surgery in Mbarara and he is in 3^rd^ year so doctors went to school just because of the outbreak”*. **–** IDI002-HSP

The outbreak also led to healthier decisions by individuals:

> *“When I fell sick of Ebola I realized that anytime I could die. I realized that now I need to plan for my future because I reached the extent of death. I used to take alcohol before Ebola but after I returned from hospital, after my dad died I sat down and asked myself, nobody gave me medicine to leave alcohol. I asked myself a question, “I have been taking this alcohol for long, what Have I benefited?”. I replied myself and decided to quit alcohol.”* – IDI004-Survivor

### Subtheme 3c: The unanswered questions

Most of the participants had vivid memories of the situation during the outbreak and had questions regarding the viral disease emphasizing the need for more research.

### Are survivors immune to Ebola?

Survivors expressed desire to know whether they had acquired natural immunity and were safe to participate in helping others in case of a future outbreak

> *“Like me I would have loved that when you do such research to find out that among us who suffered from Ebola, can’t we catch it again? I want you to do research in that area because another disaster might happen and we go there saying we survived let’s help others & end up getting the disease again. Do research on that if a person who suffered Ebola cannot catch the disease again. Do research on that.”* **–** IDI006-Survivor

### Where and how did Ebola start?

To many people this was indeed strange given that the community had never experienced such an event before. Some believed the disease’s origin to be a hunting community near the forest, since all affected members from a family living near the forest perished. A competing theory was that a traditional healer worked with a patient from Congo. However, there is still confusion as to where and how the disease started, resulting in fear that the disease could strike the community again.

> *“It has been difficult to establish the origin but it started in Kasitu Sub county currently Ngamba Sub county. There are many versions about the origin. Some people say there is a family that ate a monkey and another story in Ngamba then was that a family where the family leader is a traditional healer had a patient who came from Congo and was managed and died.”* -IDI001-HSP
>
> *“Some people ate bush meat but these are all speculations…They used to tell us during their campaigns here that Ebola is not airborne and I think that is what they say. Isn’t it? That you will not get it by someone sneezing on you…you get by contact with someone body fluids…. they just think …”* – IDI002-HSP

Several years since the outbreak, most people have generally returned to normal life except those that lost family members and the survivors. Ebola is now as well-known as malaria. When asked what they feel when they hear the word ‘Ebola,’ responses varied from that of no fear to fearful expressions with emotions associated with loss of many people. Participants also had varied reactions to the potential of another outbreak in the future.

> *“I would not fear Ebola so much, no. Because I have been there and I have seen but I fear for people who don’t know how the disease spreads.”* – IDI006-Survivor
>
> *“I feel shocked, it’s like death, that people will die again the way it happened, it was not an easy situation. It is not easy disease” and to others “A dragon monster again because of all the people who have come for research, they have told us that they have not come up with medicine to treat Ebola. So, it is a scaring word once you hear Ebola then that’s a scaring word because it’s something that cannot be treated.”* –IDI013-Administrator & affected family

To some this is the worst experience they have ever gone through and would not want to go through the same experience ever again.

> *“This time if they say that Ebola has come, I will leave boda boda business, eeeh! The truth is that the disease is dangerous. Every part of the body blood is coming out eeh! They give glove to touch someone, I still fear the disease. I saw the other man the brother to … who died in Ngamba, every part of his body was bleeding but the brother survived he works at the kikyo health center. I carried him from kikyo health center after being referred Bundibugyo.”* **–** IDI015-Boda boda rider

## Discussion

Over ten years down the line, communities in Bundibugyo still live with the experience of the 2007 Ebola outbreak that occurred in their area. This study examined the socio-cultural and economic effects of an Ebola outbreak on survivors, affected families and communities in Bundibugyo and at the time of the study, the outbreak in West Africa was on-going. Three major themes emerged during data analysis; i) The Ebola outbreak, ii) The effects of the outbreak iii) Life after the outbreak.

Host–parasite interactions are a key driver of infectious disease emergence and transmission. For human hosts these interactions transcend biological factors to include the social cultural environment. Ebola is a disruptive disease that occurs at the intersection of culture and public health[36]. Its therefore important that we include social science approaches to explain the cultural, social and the economic impacts of such outbreaks. Studies have shown that there are many social, cultural and economic changes that occur during and after the outbreak placing a significant burden on the lives of affected people [8,35] Although this outbreak was caused by a mild strain[37], the community had never witnessed people dying in large numbers before. The community response in this outbreak was driven by fear, shaping the response mechanisms before and after the confirmation of the outbreak. Fear, panic and anxiety in the community was attributed to the overall crisis communication management that was used, initially inadequate due to the strange nature of disease as studies have indicated [38]. This affected the social cultural fabric and by extension the disease dynamics particularly during the outbreak. There is fear that communities in Bundibugyo continue to live with, the fear that the disease could strike again, pushing them to continually seek for a proper answer to what caused the disease. This experience is similar to what has been described in the Kibale Ebola outbreak, where constant fear and lingering pain of the experience has been described[39]. Therefore this calls for the long-term investment in research and engagement with communities to try and understand the origin of the disease, factors that could potentially lead to outbreaks and how best to support long term community recovery. However, just like in many other Ebola outbreaks in West Africa, DRC including the most recent outbreak in Uganda, the origin remains unknown [24,40].

Several studies have linked hunting and bushmeat consumption to Ebola outbreaks [41,42]. Human contact with wildlife species, both direct and indirect, is undoubtedly an important factor in the transmission and emergence of new human pathogens from wildlife [36]. This contact is further exacerbated by the conflict with the land tenure system[43,44]. However, communities in Bundibugyo continue to hunt, eat, and trade in bushmeat and have highlighted several health and economic benefits that accrue from this practice. This finding is similar to findings in Western Uganda [45]. This is further highlighted by rejection of messaging that unilaterally stressed the health risk posed by wild meat that contradicted the experiences of target publics in West Africa Ebola outbreak, who consume wild meat without incident [41]. The communities in Bundibugyo are asking for explanations as to why hunting and bushmeat consumption are risk factors for spillover now and not before, since these are practices that have been going on for generations. It’s important to note that risk perception among such communities remains low [46], this this calls for concerted efforts among stakeholders to involve communities in the management of zoonotic diseases for early detection of such outbreaks [47].

Outbreak detection can also be enhanced by availability of in country diagnostic facilities, a key response function of the healthcare system. In the Bundibugyo outbreak, Ebola samples were tested in the USA and so what was identified as a strange disease was treated as malaria because of similarity in the presentation of the disease until confirmatory diagnosis was arrived at [48,49]. Communities also tried to seek for alternative treatment for the strange disease using witchcraft and acts of sacrifice were sighted. Tribal conflicts also played a role as Bakonzo accused the Bamba of poisoning their water source. However, these (witchcraft, acts of sacrifice and tribal accusation) resolved when the Ebola outbreak was confirmed and communities united to confront a common enemy, Ebola. This is contrary to what has been reported in the West Africa Ebola outbreak, where there was conflict between health workers and the community due to violation of cultural beliefs that are usually contradictory to prevention measures, demonstrating the value of continuous community engagement in the management of such outbreaks [50]. Lessons from COVID-19 further indicate that, heavy-handed bans risk numerous unintended consequences and tend to fail if they lack legitimacy or clash with people’s values [51].

During the outbreak, communities’ response mechanisms also involved consumption of alcohol and jik to prevent the disease. The consumption of alcohol for example was supported by community observation that some survivors were alcohol users and most of those that died, were non-alcohol users because of their religious belief. Communities believed that alcohol was a cleanser for the “Ebola germ” and offered protection to consumers. There were reports of people sustaining serious burns from jik and although many participants believed the consumption of alcohol was helpful in preventing the disease, it remains a speculation. This highlights ongoing community learning process with detrimental or beneficial effects, that is rarely documented in such outbreaks. It also highlights opportunity for partnership between local communities and use of their local observations that could be a source of insight into protective or beneficial strategies that can be taken advantage of in the battle against such emerging infectious diseases [44]. For example, in most rural communities’ consumption of alcohol is accompanied by copious consumption of meat as well [52]. Perhaps the differential health outcomes between the alcohol and non-alcohol users who are exposed to the Ebola virus could be tied to this varied access to protein between the two groups.

Overall, the effects on survivors, affected families and community varied from side effects, a sense of loss, physical and psychosocial effects among survivors and deepening tribal conflict persisting for years in some cases as already found [30]. Many survivors reported some positive outcomes such as improvement in personal hygiene and protection practices, self-awareness and health seeking behavior. However, the negative impacts were more. They include self-isolation, stigmatization. shame and embarrassment and loss of close relatives similar to what has been reported in West Africa and DRC [32]. Most significant is the impact on widows who lost their husbands. The young widows felt a great sense of loss and burden to support their children to be able to go through school. On the other hand, the older widows were haunted by the fact that their spouses did not get a decent burial and many reported withdrawing from community related activities (Self-isolation). Disease response measures can interrupt education in the short and long term. During the 2014 Ebola epidemic in West Africa and the 2018 Ebola outbreak in DRC, schools were closed, or parents were reluctant to send their children to school, due to fear of contagion, Additionally, widowed women after the loss of their spouses….struggled to take care of their children’s school needs, widows were also barred from ‘accessing their deceased husband’s land because of discriminatory inheritance laws [53], further revealing the pernicious long term effect of the disease. Effects that transcend the epidemiological triad and have far-reaching negative impacts on the social fabric of these communities.

## Conclusions

Our study has highlighted important insights into the silent epidemic of stigma and social disruption that persists long after the last contact has been traced and the infectious force of the epidemic died down. Through our focused ethnography we have shown the important role social sciences play in responding to both the short- and long-term needs of communities hit by these emerging infectious diseases. Our findings can inform future Ebola response and recovery interventions, particularly those targeting community re-integration and a mitigation of the fear and stigma associated with survivorship.

## Authors’ Contribution

^&^These authors contributed equally to this work.

## Data Availability

All relevant data are within the manuscript and its Supporting Information files.

## Acknowledgement

This project was funded by University of Minnesota as part of the capacity building under the one health work force and I would like to thank Associate Professor Katey Pelican and Chery Robertson for all the support and time they have invested in us and for maturing us into the scientists that we are today.

---

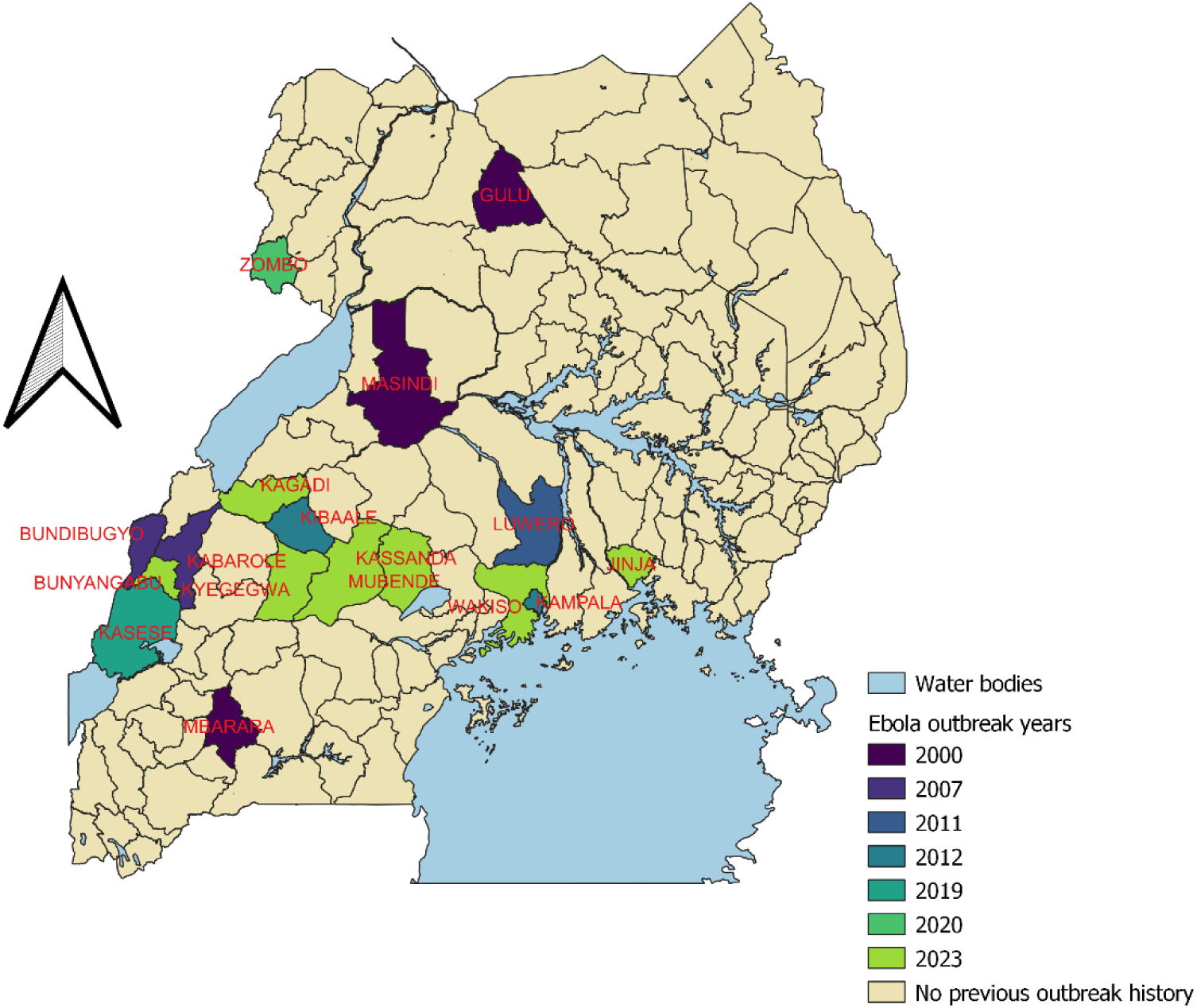

